# Offering an Internet Survey Response Mode in a Flint Water Crisis Medicaid Population

**DOI:** 10.1101/2020.11.22.20236497

**Authors:** Sabrina Ford, Kathleen Oberst, Joan Ilardo, Hong Su An, Nicole Jones, Hyokyoung Hong, Karen Clark, Zhehui Luo

## Abstract

**Objective:** To examine the preferred response mode (internet, phone, paper)to a Medicaid enrollee survey.

**Data Source:** Data was obtained from survey responses for a sub-sample of Flint Water Crisis Medicaid Expansion Waiver enrollees (N=2584).

**Study Design:** Enrollees were offered the choice of utilizing the internet, telephone, or mail to respond to a survey evaluating health services. Analyses were stratified by age, residency, race, and income. Chi-square was utilized to detect categorical differences.

**Principal Findings:** The majority (p<0.01) of participants responded by internet (55.46%), followed by mail (39.36%), and telephone (5.19%). Of those responding by internet, 75% used smart phones for connectivity. Weighted participation estimates for available survey modes showed variation by age, residence, race/ethnicity and poverty status. A smaller proportion (p<0.01) of ethnicities classified as Hispanic and Other used telephone participation compared to White or Black respondents.

Respondents at 200% Federal Poverty Level (FPL) or higher used internet at greater rates than those below 200% FPL (p<0.01).

**Conclusions:** Overall, this low-income population preferred the web-based response mode compared to paper or telephone, the variation by race and FPL suggests the continued presence of a digital divide in the population. Nevertheless, our findings revealed greater utilization of web-based tools for survey administration in vulnerable populations than was expected. This is a valuable finding that may inform future health programming and telehealth efforts particularly in the advent of COVID-19.

**What this study adds:**

- Active health care participation using internet connectivity is increasing and may be associated with improved quality and costs of health care services yet access and usage vary among populations.
- Research suggests that persons of low income and resources are unable to participate with internet based activities due to access and literacy issues that may further exacerbate health disparities.
- This study revealed enrollees of the Flint Michigan Section 1115 Demonstration utilized the internet for survey participation at a greater rate than responding by mail or phone.
- Finding that vulnerable populations have greater access to the internet via digital technology than assumed may expand opportunities to participate in their health care through health information portals and telehealth.

## Introduction/Background

Recent studies suggest low-income populations do not use or have limited access to the internet compared to higher income populations.^1-2^ Approximately 30% of non-elderly, non-dually insured Medicaid adults have never used a computer or the internet and 40% do not use email.^3^ Low computer literacy, unaffordable internet services, and lack of access to internet ready devices (e.g. mobile phones, computers, tablets, etc.) have been suggested as reasons for this observation.^4-6^ Reports indicate those who make less than $30,000 equating to over 230% Federal Poverty Level(FPL) per year are less likely to adopt or have access to internet technology.^1,7^ Thus, individuals having fewer economic resources are believed to have low access to internet services and equipment.

Federal programs have been implemented to address the disparity in internet access. Among these are ConnectHomeUSA,^8^ which is intended to provide internet accessibility in public spaces and low-income housing. In addition, the Federal Communications Commission (FCC) Lifeline Support for Affordable Communications (Lifeline) program offers reduced fees for mobile phones and internet services for individuals at or below 135% of the poverty level.^9,10^ This includes individuals and families who qualify for Medicaid, Supplemental Nutrition Assistance Program (SNAP), Supplemental Security Income (SSI), and/or Public Housing Agency (PHA) programs.^10-12^ While these programs address some access issues, barriers still exist in the form of internet related literacy and affordability of internet ready devices.^13^

A study conducted by Deloitte^14^ found that Medicaid enrollees have increased accessibility to smart phones and tablets, but at a significantly lower rate than employed medically insured persons. Likewise, fewer low-income families have access to personal computers in the home, and instead, primarily use smart phones to access the internet compared to those with middle and high incomes.^15^ The coronavirus pandemic (COVID-19)has made this digital divide even more stark.^16,17^ This continued disparity can be problematic for low-income patients accessing healthcare portals that are optimized for computer monitor screens and in turn, can limit usability for people who do not own a computer.^18^ Few empirical and healthcare quality studies examine Medicaid enrollees’ access to the internet and related devices and how they utilize the service. Furthermore, academic reports on technology use for personal health quickly become outdated because of frequent changes in computer technology.^1,19^

Internet service now provides access to a variety of tasks formerly conducted via telephone or paper. In particular, the ability to administer surveys online provides individuals with means to participate at their convenience in a secure, anonymous manner. Survey platforms such as SurveyMonkey (SurveyMonkey Inc.) or Qualtrics, (Qualtrics Inc.) can accommodate participation validation methods as well as allow participants to complete a survey in more than one sitting resulting in improved response rates. These platforms further provide a seamless approach to ensure participants accurately follow complex skip patterns and jumps based on key responses. Thus, internet-based surveys allow for increased completion rates and minimize the occurrence of invalid responses.^20^ Another benefit to online survey methodology is the immediate availability of analyzable datasets. Online survey participants contribute not only to direct data entry but reducing delay from data collection to analysis. Due to the minimization of subsequent data entry and the need for data entry validation, online survey processes are more resource efficient. This rapid turnaround of findings may support more timely program evaluation efforts and facilitate more real-time assessments of program modifications.^20-21^

We planned an enrollee survey as a component of a Medicaid Waiver authorization evaluation. Adding an online response method provided the evaluation team with an opportunity to examine response rates of different participation modes. Based on current reports of lower internet use in low-income Medicaid populations, we expected internet survey responses would be lower than mail or telephone participation rates. Further, we were interested to identify how differences in response mode varied by race/ethnicity, income, and geographic residency.

## Survey Data and Methods

A description of activities for the 1115 Waiver Evaluation plan was submitted to the Michigan State University (MSU) Human Research Protection Program (HRPP) and determined not to be research involving human subjects. All communications including enrollee letters, reminder postcards and surveys were submitted to the Michigan Department Health and Human Services (MDHHS) for review and approval.

The Center for Medicare and Medicaid Services (CMS) funded the Flint 1115 Medicaid Expansion Waiver that was implemented in 2016. The waiver offers additional family support coordination services for pregnant women and children exposed to lead during the Flint Water Crisis. In addition to expanded healthcare services, the qualifying income increased from at or below 133% of the FPL to 400% FPL. Individuals above 400% FPL may also enroll in the waiver through cost-sharing.

The 1115 waiver authorization required a formal evaluation of the success to which key objectives were achieved. The Institute for Health Policy (IHP) at Michigan State University managed the evaluation process. One component of the evaluation plan was to conduct enrollee surveys to gather information about the utilization of and satisfaction with waiver services and participation. The survey also served to provide an opportunity for participants to report on changes in health outcomes attributed to waiver enrollment.

### Survey Design and Procedures

Hypotheses included in the approved evaluation proposal guided the enrollee survey design and question development. Separate yet similar surveys were designed to address child and adult outcomes. The child version included additional questions pertaining to developmental and educational factors known to be related to lead exposure. The child version of the survey was distributed to parents or guardians of enrollees 0-17 years of age at the time of enrollment. National survey questions regarding self-reported health status were included in the surveys. An IHP evaluator with experience in psychometric testing and survey design formulated initial questions for topics where no nationally accepted questions were identified. The surveys were reviewed by the full evaluation team and survey experts affiliated with the Office for Survey Research (OSR) at MSU’s Institute for Public Policy and Social Research.

Due to concerns about internet accessibility, the evaluation team originally developed a plan including only telephone and paper response options. Prior to survey implementation, the evaluation team sought community feedback from two groups: A Flint Area Community Advisory Board and the Michigan State University-Hurley Children’s Hospital Pediatric Public Health Initiative Parent Partners. The Parent Partners group includes fifteen representatives from all nine wards of the City of Flint as well as parents from the Greater Flint area. Group members included parents, foster parents, grandparents and guardians. Key feedback from the group included input on survey content, language for instructions, and delivery mechanisms. Particularly, members suggested a web-based survey participation option accessible by smart phone in addition to the proposed telephone and mail options.

The study design included three waves of enrollee surveys occurring at nine-month intervals. Wave 1 represented baseline measurements with Wave 2 (9-month) and Wave 3 (18-month) follow-up points planned. Based on the Flint residents’ feedback, revised survey mode options included 1) web-based participation using two factor authentication, 2) telephone call-in to the survey center, and 3) postal mail option for paper surveys. This report describes response rates by mode for the Wave 1 survey.

To increase the odds of internet response a short bitly web address and assigned study IDs and passcodes were utilized. This information was disseminated to individuals selected for the sample through the mailed introductory letters. The baseline survey provided an opportunity for individuals to enter an email address and telephone number with permission to contact them by SMS so that invitations to upcoming survey waves could be more efficiently directed by the online survey system.

### Enrollee Sampling Method

The sampling pool (N = 24,082) included all waiver enrollees having at least six months continuous enrollment at the time of the survey (i.e., the target population). The data for the sampling pool was acquired from the MDHHS Data Warehouse compliant with MSU HRPP. Stratified random sampling was used to select the survey sample. Table 1 shows the sampling proportions by strata.

**Table 1.** Characteristics of All Eligible Enrollees and Selected Survey Participants

| Category | All Waiver Enrollees (N=24,082) | Selected Sample (N=11,453) | p-value |
| --- | --- | --- | --- |
| Age Class |  |  | 0.29 |
| 0 - 6 years | 8,675 (36.2) | 4,126 (36.0) |  |
| 7 - 17 years | 13,023 (54.1) | 6,158 (53.8) |  |
| 18 or older | 2,384 (9.9) | 1,169 (10.2) |  |
| Residence Class |  |  | <0.01 |
| Always in Genesee | 21,584 (89.6) | 10,068 (87.9) |  |
| In and Out of Genesee | 384 (1.6) | 384 (3.4) |  |
| Never in Genesee | 2,114 (8.8) | 1,001 (8.7) |  |
| Race/ Ethnicity |  |  | 0.63 |
| Non-Hispanic White | 7,923 (32.9) | 3,756 (32.8) |  |
| Non-Hispanic Black | 14,352 (59.6) | 6,822 (59.6) |  |
| Hispanic | 830 (3.4) | 413 (3.6) |  |
| Other/Unknown | 977 (4.1) | 462 (4.0) |  |
| Federal Poverty Level (%) |  |  | 0.51 |
| 0 - 99% | 12,204 (50.7) | 5,843 (51.0) |  |
| 100 - 199% | 9,422 (39.1) | 4,437 (38.7) |  |
| 200% and above | 2,456 (10.2) | 1,173 (10.2) |  |

Enrollee age class, Genesee County residency, race/ethnicity, and FPL as documented in the Data Warehouse administrative files were used as strata. Three age groups were established: 0-6 years, 7-17 years, and 18+ years of age at the time of waiver enrollment. The residency category included three groups: Always lived in Genesee County, Lived In & Out of Genesee County and Never lived in Genesee County. Individuals exposed to the Flint Water did not necessarily live within the City of Flint or in Genesee County. Exposures through employers, daycare or schools, and healthcare facility location(s) were documented. In recognition of the potential for geographic dispersion, the waiver further permitted ongoing coverage for individuals relocating outside of the greater Flint area after documented exposures. Race/Ethnicity strata included non-Hispanic White, non-Hispanic Black, Hispanic and Other categories. FPL included three strata of up to 99% FPL, 100-199% FPL and 200% or above.

Due to the longitudinal nature of documenting health outcomes and concerns over participant attrition, the sampling rate was 50% for the first survey wave. However, for some categories where small numbers of enrollees were documented, all eligible persons were included in the sample. Over 11,000 (n=11,453) enrollees were selected for inclusion into the Wave 1 survey. Table 1 displays the characteristics of total eligible enrollees by strata and the count of selected sample individuals. Generally, the characteristics of the selected sample reflect the population except for residence class where over-sampling was intentional due to small cell size.

Another concern leading to the large sampling frame was the national interest in conducting research on persons in the region after the water crisis. Many research projects were simultaneously recruiting community participation and the success of encouraging participation was worrisome. This also led the evaluation team to secure approval to provide small monetary incentives of $10 per completed survey to promote participation. The incentive payments were structured so that if a participant submitted all three survey waves, they would receive a $20 bonus payment.

After sample selection, the MDHHS Data Warehouse was accessed to extract most currently available enrollee mailing information. An honest broker approach was used to obtain the identifiable data necessary to execute the mailing. This identifiable data file was hand-delivered on an encrypted and password protected external drive to the OSR for survey implementation. Survey response data was returned to the evaluation team with a random survey ID assigned by OSR.

The survey was programmed into Qualtrics (Qualtrics Inc.) for the web-based option. Qualtrics is an online survey platform that organizes data intended for statistical analyses. A short web address was created to reduce the keystrokes required of participants to reach the survey page. Access to the survey was controlled using the unique, single-use study ID to prevent participants from completing the survey more than once. The online survey was further protected from non-waiver enrollee participation by the use of two-factor authentication along with restrictions imposed on the ability of internet search engines to locate the survey. All survey materials regardless of mode were provided in English.

OSR used a batch number procedure to manage the mailing process due to the large sample. The sampled enrollees were randomly divided into four batches to track and expedite the mailing process. Each batch included 2,863 individuals, except for batch four, which included 2,864 totaling 11,453.

Preliminary notification letters were mailed for each batch and included instructions on available modes by which to complete the survey and information regarding the participation incentive. These letters included the individual’s assigned study ID with instructions on accessing the web-based survey system and toll-free telephone numbers to call-in to the survey center in order to complete the survey. Participants were informed they would receive a paper version if no response was recorded through internet or telephone after two weeks.

If no response was documented within two weeks of the initial mailing, a second mailing with a paper copy of the survey was disseminated through the postal service using the same batch assignment. The cover letter accompanying the survey also informed the participant about the alternative options of telephone or web-based participation. A third mailing was sent after three weeks in the event no response was recorded. This letter again reminded individuals of the telephone and web options and invited individuals to request another paper version if they had misplaced the one originally sent. Finally, a fourth reminder letter was mailed to all non-responders encouraging them to participate.

### Data Collection

All paper surveys were entered by OSR staff using a blinded, double data entry process. Surveys completed by telephone were monitored by OSR supervisory staff with data entry managed through a Computer Assisted Telephone Interview (CATI) system. Web-based survey responses were directly entered by participants and captured by Qualtrics software in a format readily exportable to spreadsheet and statistical software. All forms of data were compiled and cleaned using SAS 9.4 software (SAS Institute, Inc.)

### Statistical Analyses

The randomness of the survey sample was assessed by comparing the population to the sampled survey participants by age class, residence class, race/ethnicity, and FPL. Response rate was also compared by participant’s characteristics using chi-square test. The association between the respondent’s characteristics and the response mode was tested using a chi-square test. In the case of small cell size less than 5, we used the exacted p-value estimated by Monte Carlo simulation. All chi-square and exact tests were performed by SAS 9.4 software.

## Study Results

The selected survey sample was representative of the population as presented in Table 1. There were no significant differences of proportion between the eligible population and selected sample for age class, race/ethnicity, and FPL. Because the residence category of *In and Out of Genesee County* contained few individuals, we oversampled this group. Thus, there was a statistically significant (p<0.01) higher proportion of this class in the selected sample.

Of the 11,453 survey invitations mailed across the four batches, 2584 (22.6%) of participants responded (Table 2). There was no mailing batch effect for response (data not shown, p=0.07) therefore all responses were combined to create a single cohort of respondents for analysis. Of the 2584 returned surveys, 2359 (91.3%) were child enrollees and 225 (8.7%) reflected adult enrollees. After data cleaning, 2356 (99.8%) of the child surveys and all the adult surveys were retained for reporting.

**Table 2.** Comparison of Selected Sample by Survey Response Category

| Category | Survey Response |  | p-value |
| --- | --- | --- | --- |
|  | No<br>N=8,869 | Yes<br>N=2,584 |  |
| Age Class |  |  | <0.05 |
| 0 - 6 years | 3,199 (36.1) | 927 (35.9) |  |
| 7 - 17 years | 4,726 (53.3) | 1,432 (55.4) |  |
| 18 or older | 944 (10.6) | 225 (8.7) |  |
| Residence Class |  |  | 0.67 |
| Always in Genesee | 7,786 (87.8) | 2,282 (88.3) |  |
| In and Out of Genesee | 304 (3.4) | 80 (3.1) |  |
| Never in Genesee | 779 (8.8) | 222 (8.6) |  |
| Race/ Ethnicity |  |  | <0.01 |
| Non-Hispanic White | 2,804 (31.6) | 952 (36.8) |  |
| Non-Hispanic Black | 5,386 (60.7) | 1,436 (55.6) |  |
| Hispanic | 306 (3.5) | 107 (4.1) |  |
| Other/Unknown | 373 (4.2) | 89 (3.4) |  |
| Federal Poverty Level (%) |  |  | <0.01 |
| 0 - 99% | 4,544 (51.2) | 1,299 (50.3) |  |
| 100 - 199% | 3,469 (39.1) | 968 (37.5) |  |
| 200% and above | 856 (9.7) | 317 (12.3) |  |

Table 2 compares the survey respondents to the non-responders. Chi-square tests revealed significant differences in age groups, race/ethnicity and poverty level between these groups. Responders were more likely to reflect enrollees between 7-17 years of age, White and over 200% FPL. Eligible Black participants had significantly lower participation than other race groups except the Other/unknown group (p<0.01). There was no statistically significant difference between White and Hispanic groups.

## Demographic Characteristics and Survey Response Mode

Since there were significant differences in response rates by demographic characteristics for age, race, and FPL, weights were used to adjust for sampling bias. Table 3 presents associations between population characteristics and response mode based on the adjusted weight. The overall proportion of online participation exceeded other survey modes among all responders. More than half (55.6%) completed the survey using the internet method. Nearly 40% (39.3%) participated using the postal mail option while just 5.2% used the telephone option. Significant differences in response mode were observed for all demographic characteristics. Regardless of demographic characteristics, internet response showed the highest rate compared to paper or telephone.

**Table 3.** Weighted Population Characteristics by Response Mode

| Characteristics | Response Mode |  |  | Total<br>N=24,802 | p-value |
| --- | --- | --- | --- | --- | --- |
|  | Internet<br>(55.6) | Paper<br>(39.3) | Phone<br>(5.2) |  |  |
| Age Class |  |  |  |  | <0.01 |
| 0 - 6 years | 5175 (59.7) | 3032 (35) | 468 (5.4) | 8,675 |  |
| 7 - 17 years | 6739 (51.7) | 5584 (42.9) | 700 (5.4) | 13,023 |  |
| 18 or older | 1473 (61.8) | 837 (35.1) | 74 (3.1) | 2,384 |  |
| Residence Class |  |  |  |  | <0.01 |
| Always in Genesee | 12031 (55.7) | 8371 (38.8) | 1182 (5.5) | 21,584 |  |
| In and Out of Genesee | 197 (51.3) | 182 (47.5) | 5 (1.3) | 384 |  |
| Never in Genesee | 1143 (54.1) | 895 (42.3) | 76 (3.6) | 2,114 |  |
| Race/Ethnicity |  |  |  |  | <0.01 |
| Non-Hispanic White | 4344 (54.8) | 3387 (42.8) | 191 (2.4) | 7,923 |  |
| Non-Hispanic Black | 8125 (56.6) | 5167 (36) | 1059 (7.4) | 14,352 |  |
| Hispanic | 427 (51.4) | 396 (47.7) | 8 (0.9) | 830 |  |
| Other/Unknown | 472 (48.3) | 461 (47.2) | 44 (4.5) | 977 |  |
| Federal Poverty Level |  |  |  |  | <0.01 |
| 0 - 99% | 6404 (52.7) | 4878 (40.1) | 880 (7.2) | 12,161 |  |
| 100 - 199% | 5122 (56.8) | 3548 (39.4) | 346 (3.8) | 9,016 |  |
| 200% and above | 1843 (63.4) | 986 (33.9) | 77 (2.6) | 2,905 |  |

In the case of age class, adults (61.8%) showed the highest rate of internet response with nearly a 10% difference between adult and 7-17 years group. With respect to residence class, the *In and out of Genesee* group had significantly different response modes compared with *Always in Genesee* and *Never in Genesee*. The use of internet and paper response among the *In and Out of Genesee* was similar with less than 5% difference. Chi-square revealed these categorical differences a significant (p<0.01). Black enrollees had higher response rates using internet and telephone and were less likely overall to respond by paper than individuals in other categories. Conversely, individuals classified as Hispanic and Other/Unknown race were more likely to respond by paper than White or Black individuals (p<0.01). Figure 1 presents response rate by race/ethnicity. The internet response rate increased with increasing FPL (Figure 2). However, regardless of FPL level, the internet survey was the preferred response mode. More than 50% used the internet survey for the less 99% FPL group. For FPL 200% or above group, 63.4% used the online survey compare to the paper survey (33.9%) or the telephone (2.6%).

**Figure 1.**
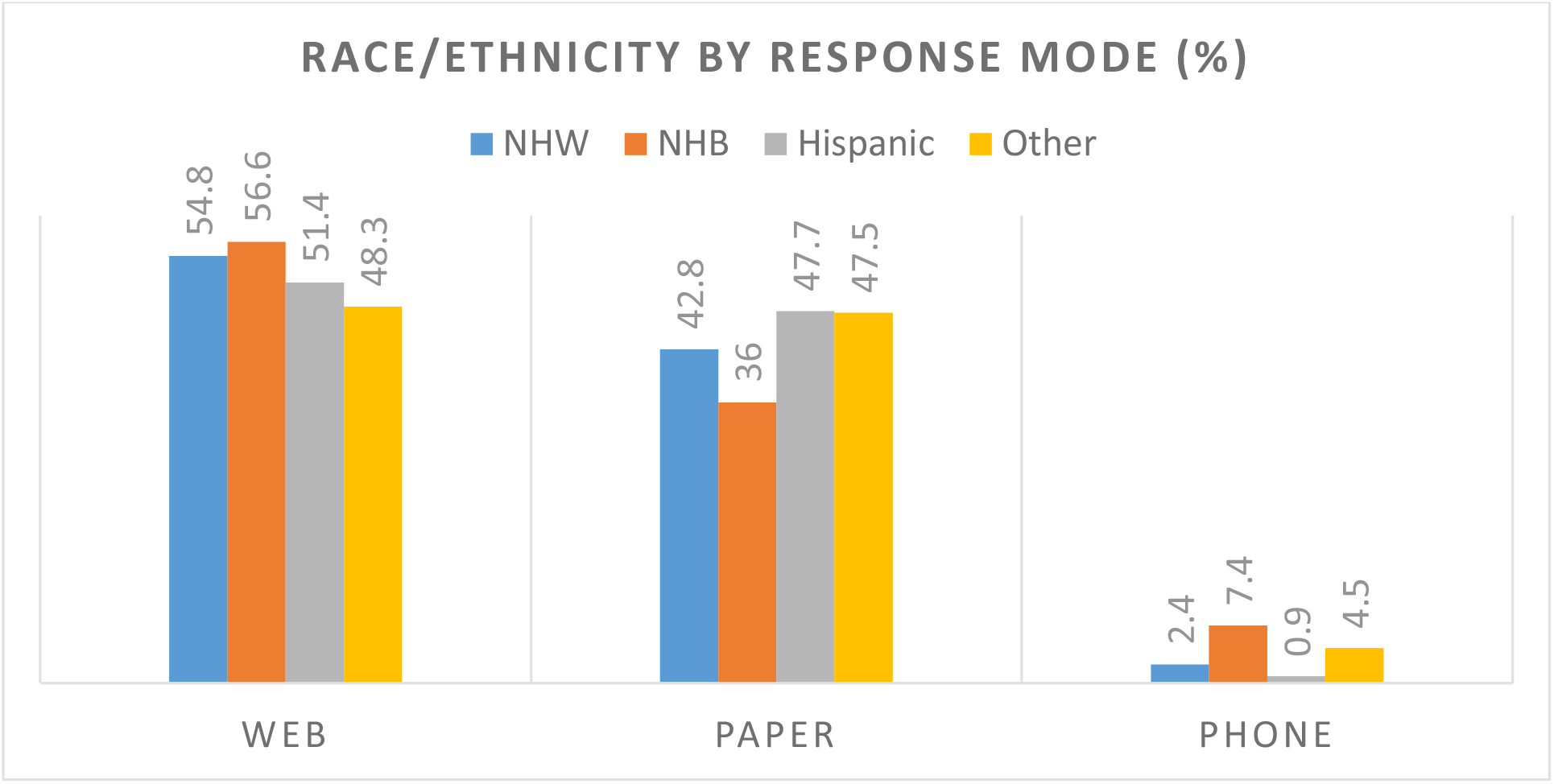
Race and Ethnicity by Type of Response to Survey.

**Figure 2.**
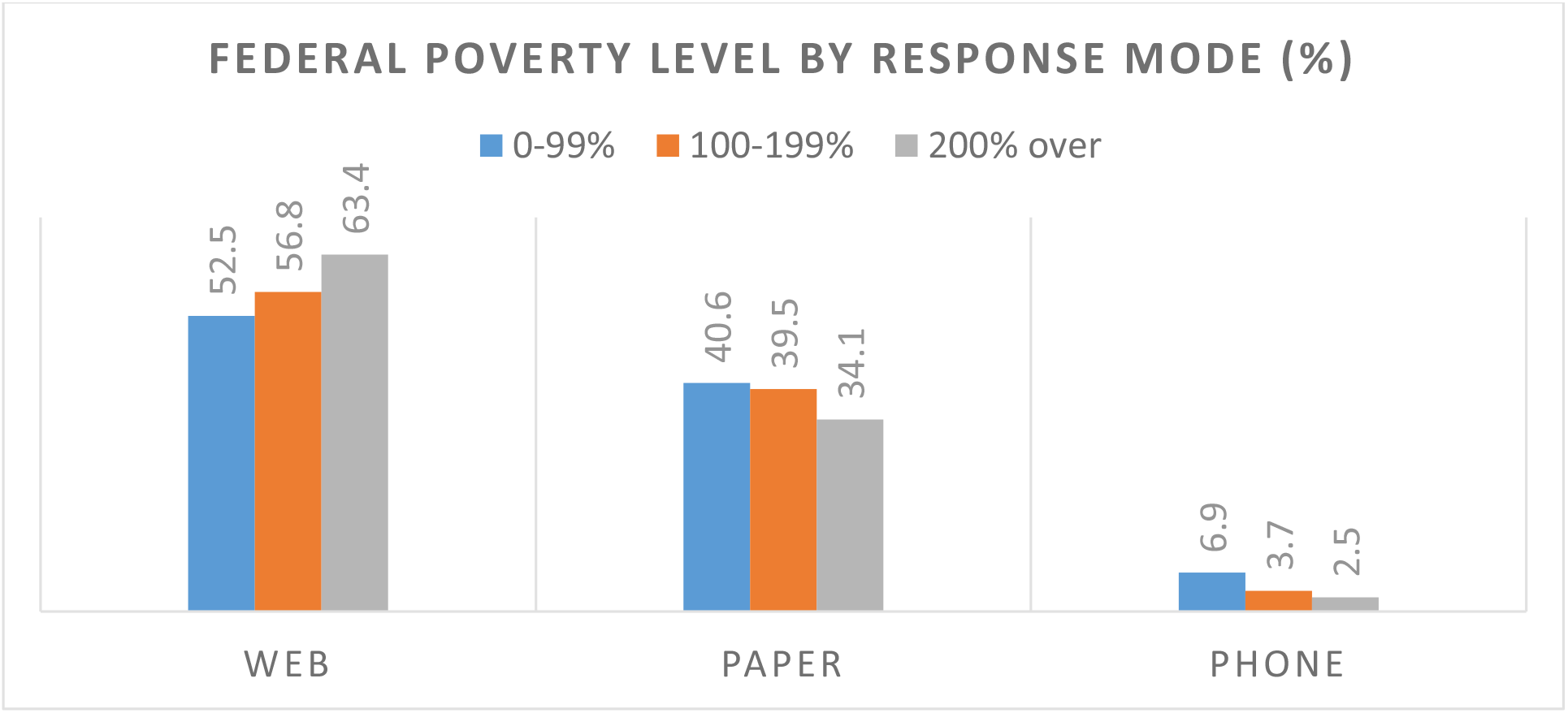
Federal Poverty Level by Type of Response to Survey.

An operational benefit from internet participation was that over 70% of respondents provided email addresses and/or mobile numbers to receive email or text messaging to notify them of future survey waves.

## Discussion

In this report, we documented higher participation with a web-based survey compared to paper or telephone options. Respondents were drawn from enrollees of the Flint Water Medicaid Expansion Waiver. The ordered preference for survey participation method was consistent regardless of race/ethnicity, residence category, or FPL of selected enrollees. Of the 2,584 respondents, the majority (55.5%) responded using the internet option, followed by mailed paper surveys (39.4%) with fewer respondents (5.2%) choosing the telephone option.

While the greatest proportion of respondents within each race/ethnicity category used the online option, the rates of postal mail versus telephone option did vary. Of note, those who were identified as Hispanic or categorized as “Other/Unknown” based on the MDHHS administrative enrollment data were significantly less likely to use the telephone option compared to Black or White. This may reflect the availability of and or comfort with conducting the survey using English as a spoken language. Those having English as a second language may prefer options that permit them more time to look up unfamiliar words and/or take advantage of friends, family or other tools to facilitate translation. Unfortunately, since this data was not collected, we cannot ascertain this premise.

This waiver allowed those at higher income levels (>212% FPL) to enroll without cost. Even individuals exposed to the Flint water exceeding 400% FPL were eligible to enroll by paying a monthly fee. This enabled the evaluation team to assess for participation differences among varying income levels. Across all income levels, the internet participation mode was most prevalent.

Prevailing opinions are that accessible survey methods for a vulnerable population include only paper and telephone.^1-3^ Thus, inclusion of web-based survey methods are generally overlooked due to perceptions regarding lack of access to internet services and technology. This evaluation team’s original survey plans followed prevailing opinion. However, input from community advisory groups recommended the evaluation team consider internet and smart phone technology as a means to access the survey. The recommendation proved fruitful.

In addition to concerns regarding internet access, another potential barrier identified with an online survey process was the lack of email addresses that are used by these systems to deliver invitations and participation links. Despite not having email addresses for enrollees for Wave 1, the survey coordinators used the available postal contact information to distribute web address information and found that many respondents used the web-based survey as well. The finding that 70% offered their email address for follow-up waves of the survey revealed access and willingness to use digital communications.

Among the majority who completed the online version of the survey, we documented 75% using smart phones as the means of access. Since most of the respondents who accessed the internet using smart phone technology there is great the potential to address health status and disparities using digital platforms.^15^ In addition, smart phone access can mitigate the need for more costly in-home personal computer and internet service for persons having low-income.

Investing in the tools and education to increase the opportunity to use internet accessible technology would not only support health equity but also improve other facets of life. A significant amount of societal involvement involves internet technology. This has become apparent during the COVID-19 pandemic as schools, health care, human services and businesses have shifted from in-person interactions to telehealth and social networking platforms.^22-23^ The online survey participation experience of this cohort of Medicaid enrollees demonstrates potential to address other health needs leveraging digital technologies. Current studies have not emphasized the ability of those with fewer economic resources to participate in informing service delivery improvement using digital technology. In addition to inquiries about quality or access to health care services, web-based services may further result in cost-savings. As an example, this evaluation process was able to maximize technology to reduce administrative costs associated with printing, stuffing, mailing, and manually entering survey data.

These findings are encouraging but further evidence is needed to discern associations between uses of technology and specific characteristics of medically underserved populations. These results show the degree to which this particular cohort was able to overcome the first presumed barrier of access to the internet. If a reasonable likelihood exists that access is available, these findings may support testing technology with the goal of increasing health literacy, communicating with health providers and insurers, and advocating for health concerns.

Substantial gaps will continue to exist for those Medicaid enrollees who lack direct access to or use the internet^18,19^ and we see continued evidence of the digital divide in health disparities during the COVID-19 pandemic.^16^ However, it is imperative that internet technology be made available universally for vulnerable populations to reduce health disparities and reduce administrative costs that could be re-invested into direct care. In order to fully test this, we suggest further investigation among those who still remain unable to connect and interact with technology. Solutions may include individual education about gaining access to the internet as well as social supports to address socio-economic hurdles such as subsidized smart phones or public computers and wireless connectivity.

The use of online resources also has the potential to build trust for individuals who may have difficulty communicating with their physician or disclosing adverse or risky health behaviors.^18^ Eliminating the shame and embarrassment of disclosing health concerns preliminarily via secure healthcare internet portals, may mitigate fear and discomfort of talking about health concerns when meeting health providers in person.^24,25^

### Limitations

We describe participation rates by survey mode using a natural experiment associated with implementing a survey with Flint Water Medicaid Expansion enrollees. This evaluation was not designed, nor participants randomly assigned to empirically compare the utilization of various survey response options. All selected individuals in the sample were offered an opportunity to select among all three response options (internet versus telephone versus postal mail). Individuals self-selected their preferred method.

Another limitation was the inability to determine the influence of responder age. The available age categories reflected the enrollee’s age of the enrollee rather than that of the individual completing the survey on behalf of the enrollee. This prevented the evaluation team from evaluating the impact of differences in internet use by age cohorts which documents younger individuals are more apt to utilize the internet to attend to daily activities.^26^ While it may be reasonable to expect those eighteen or older answering on their own behalf, we cannot say anything about the surveys that targeted minors as these were directed to the parent or guardian as reflected in the state’s administrative data.

## Conclusion

The evaluation process was able to take advantage of a community-based suggestion to measure the degree to which a vulnerable population would participate in an online survey. The outcomes of this experience have practical implications for further exploration of digital health equity initiatives targeting lower-income families.^22^ Use of digital health technology can inform and educate individuals who face barriers to equitable health services. The findings warrant a larger study to identify preferred mechanisms of low-income populations who use digital technology. Such a study may yield information that can be used to train and educate providers on meaningful ways to incorporate technology into their practice and increase health literacy in patients who are not currently using technology to communicate about their healthcare.

The methodological design of this study may be helpful to other entities involved in quality improvement and evaluation of health care delivery. The online participation option provided a number of benefits to the evaluation process. Specifically, the timeliness of the information was improved compared to the paper survey option. Further, complex skip patterns were embedded in the online survey and seamless to the end-user resulting in only applicable questions being asked. Additionally, fewer resources were needed to collect these data as individuals carried out their own data entry. Demonstration programs needing information in a relatively rapid timeframe or wanting to adjust programs in real-time may wish to consider this option.

Providing low-income populations with internet access options can improve their access to appropriate health services as well as provide pertinent health information and education.^27-29^ Additionally, the need to connect low-income individuals to internet technology is critical as it becomes an increasingly integral part of the fabric of our environment as demonstrated during the COVID-19 pandemic.

## Data Availability

Data used for this study were obtained from a participant survey and securely stored at Michigan State University but belong to the State of Michigan Department of Health and Human Services (MDHHS). Permission to use the data is required from the MDHHS.

